# Did COVID-19 Vaccines Go to the Whitest Neighborhoods First? Racial Inequities in Six Million Phase 1 Doses Shipped to Pennsylvania

**DOI:** 10.1101/2022.03.12.22272300

**Authors:** Geoffrey S. Holtzman, Yukun Yang, Pierce Louis

**Affiliations:** Center for Antiracist Research, Boston University; School of Public Health, Boston University

**Keywords:** COVID-19, health disparities, race, racial equity, vaccine access, vaccine allocation, vaccine hesitancy

## Abstract

Research on racial disparities in COVID-19 vaccination rates has focused primarily on vaccine hesitancy. However, vaccine hesitancy research is increasingly unable to account for racial disparities in vaccination rates in the U.S., which have shrunk rapidly over the past year. This and other evidence suggests that inequities in vaccine allocation and access may have contributed to vaccination rate disparities in the U.S. But to our knowledge, no previously published research has examined whether the geographic distribution of COVID-19 vaccines has led to greater access for White Americans than for Black Americans.

Here, we link neighborhood-level data on vaccine allocation to data on racial demographics to show that in the first 17 weeks of Pennsylvania’s COVID-19 vaccine rollout (Phase 1), White people were 25% more likely than Black people to live in neighborhoods (census tracts) that received vaccine shipments. In the 17 weeks of Pennsylvania’s *de jure* restrictions on vaccine eligibility, *de facto* geographic restrictions on vaccine access disproportionately disadvantaged Black people and favored White people. In revealing these vaccine inequities, our work builds on prior work to develop a theory-driven, evidence-based, reproducible framework for studying racial inequities in the distribution of COVID-19 vaccines.

## Introduction

Racial and ethnic disparities in COVID-19 vaccination rates in the U.S. have been widely discussed in the media and in scientific literature^1,2^. The dominant narrative is that these disparities are driven by differences in vaccine hesitancy^3–7^. That explanation is plausible, given the 400-year history of race-based medical atrocities and inequities in (and *en route* to) the Americas^8,9^. It is also supported by a great deal of survey research, which has generally found greater COVD-19 vaccine hesitancy among Black and Hispanic/Latino/a/e/x people than White people in the U.S.^1,10–12^. However, there are a number of reasons to believe that this prevailing explanation for vaccination rate disparities in the U.S. is at best incomplete.

Racial and ethnic disparities in COVID-19 vaccine hesitancy in the U.S. have declined only slightly in the past year^13,14^, whereas Black/White disparities in vaccination rates have dropped by over 90% in that time^15,16^. In at least nine states, including Pennsylvania, Black people are now more likely than White people to have been vaccinated^17^. Nationally, the Hispanic and Latino/a/e/x population has now been vaccinated at higher rates than the non-Hispanic, non-Latino/a/e/x population^15^. Racial disparities in vaccine uptake have been observed in the U.S. even among people who are not hesitant to be vaccinated^13^. In contrast, little or no racial disparity in vaccine uptake has been observed in the U.K., despite racial disparities in vaccine hesitancy in the U.K. similar to those observed in the U.S.^13^. Collectively, these findings suggest that “issues related to access may underlie the observed lower vaccine uptake among minority populations in the U.S.”^13^. In that case, new approaches are needed to help understand and overcome the barriers to racial and ethnic equity in COVID-19 vaccine access in the U.S.

One effective technique for studying health disparities is to link neighborhood–level data on the availability of scarce resources (like COVID-19 vaccines) to data on racial and ethnic demographics^18^. If demographic differences in health outcomes track geographic differences in health resource availability, racial and ethnic disparities in health outcomes may be remedied by more equitably allocating healthcare resources across neighborhoods. This technique has provided critical insights into a number of complex geodemographic relationships, illuminating some of the hidden causes of Black-White disparities in heart disease (supermarket access) and cancer (air quality)^19,20^.

Here, we draw on this methodology to analyze the relationship between the racial and ethnic populations of 3,218 neighborhoods (defined as census tracts^21^) and the shipping destinations of 6,365,810 COVID-19 vaccine doses allocated across Pennsylvania. Because our emphasis is the just distribution of scarce medical resources^22–25^, we focus on Phase 1 of Pennsylvania’s vaccine distribution plan (December 14, 2020 through April 12, 2021)^26^. During this period, the state prioritized certain groups (e.g., all adults aged 65 or older) as vaccine-eligible in order “to ensure ethical allocation of scarce vaccine”^27^. We chose Pennsylvania because it was the only U.S. state to provide separate datasets for doses allocated by the state Department of Health (DoH) and by two Federal Retail Pharmacy Partnership programs (Moderna-retail and Pfizer-retail)^26^.

For DoH, doses were allocated by the Centers for Disease Control and Prevention (CDC) to the Pennsylvania state government^28^. DoH then allocated these dose shipments to pharmacies, hospitals systems, county health departments, and other public and private entities, many of which had multiple points of service^29^. For both retail programs, the CDC allocated doses directly to corporations operating pharmacy chains in Pennsylvania^30–32^. Those corporations then determined which of their retail locations would receive vaccine shipments and how many doses those shipments would contain^33^.

With these data, we were able to test three hypotheses associated with four basic questions concerning vaccine equity:

*H*_1_: Were *Whiter neighborhoods* more likely to receive at least *some vaccine* shipments?
*H*_2_: Did *Whiter neighborhoods* receive *more doses* when they received vaccine shipments?
*H*_3_: Did *White people* tend to have *more doses* shipped to their neighborhoods overall?

By separately analyzing the DoH, Moderna-retail, and Pfizer-retail datasets, we were able explore one additional question:

*H*_4_: Did *retail* pharmacy chains allocate vaccine across their Pennsylvania stores locations in a way that was *less equitable* (with respect to *H*_1_, *H*_2_, and *H*_3_) than DoH allocations?

In other words, we were able to explore whether public-private partnerships^34^—which made rapid development and production of COVID-19 vaccines possible^35,36^—might have had negative consequences for vaccine equity^37–39^. Such explorations may prove critical in determining how to equitably distribute the variant-specific boosters now being tested in humans^40,41^.

## Results

### Statewide descriptive statistics

Table 1 provides demographic information for all neighborhoods, as well as separate demographics for areas that did and did not receive any of the 6,365,810 vaccine doses in our dataset. Neighborhoods receiving no vaccine during Phase 1 (*n* = 2,217) had an average of 32.8% more Black residents per capita (11.6%) than neighborhoods that did receive vaccine (8.8%). By contrast, we observed just 0.7% fewer adults under age 65 (who were ineligible to vaccinated at that time) in those neighborhoods (61.1% vs. 61.5%).

**Table 1:** Demographics of neighborhoods that did and did not receive vaccine shipments. Vaccine access by demographic category calculated as percentage of all Pennsylvanians of a given race, ethnicity, or age living in neighborhoods receiving vaccine shipments.

| | Neighborhoods not<br>receiving vaccine<br>($n = 2,217$) | Neighborhoods<br>receiving vaccine<br>($n = 1,001$) | All neighborhoods<br>in Pennsylvania<br>($n = 3,218$) | Vaccine access<br>by demographic<br>category |
| --- | --- | --- | --- | --- |
| **Race/ethnicity** |  |  |  |  |
| White | 6,405,016 (75.4%) | 3,372,147 (78.4%) | 9,777,163 (76.4%) | 34.5% |
| Black | 986,899 (11.6%) | 376,144 (8.8%) | 1,363,043 (10.7%) | 27.6% |
| Hispanic/Latino | 635,501 (7.5%) | 299,715 (7.0%) | 935,216 (7.3%) | 32.0% |
| Other | 465,504 (5.5%) | 250,604 (5.8%) | 716,108 (5.6%) | 35.0% |
| **Age** |  |  |  |  |
| <18 | 1,790,710 (21.1%) | 871,681 (20.3%) | 2,662,391 (20.8%) | 32.7% |
| 18-64 | 5,222,069 (61.5%) | 2,625,350 (61.1%) | 7,847,419 (61.3%) | 33.5% |
| 65+ | 1,480,141 (17.4%) | 801,579 (18.6%) | 2,281,720 (17.8%) | 35.1% |
| **TOTALS** | **8,492,920 (100%)** | **4,298,610 (100%)** | **12,791,530 (100%)** | **33.6%** |

Strikingly, White Pennsylvanians (34.5%) were 25.0% more likely than Black Pennsylvanians (27.6%) to live in neighborhoods that received vaccine shipments. Despite being universally prioritized for vaccination^29,42^, Pennsylvanians aged 65 or older were only 5.0% more likely to live in neighborhoods with vaccine (35.1%) than neighborhoods without vaccine (33.5%).

### Pfizer-retail statistical tests

The odds of a neighborhood receiving any Pfizer-retail vaccine increased by 18.1% for every 1,000 White residents (*OR* = 1.18). Among neighborhoods receiving vaccine, an additional 599 doses could be expected per 1,000 White residents (Figure 1a). Additional residents who self-identified on the Census as Hispanic or Latino or as members of other races had no significant effect on the expected odds of receiving any vaccine, nor did they have a significant effect on quantity of doses received (Table 2).

**Table 2:** Effect of White and Black neighborhood populations on allocation of any vaccine. Estimates represent change in log odds of neighborhoods (*n* = 3,218) receiving any COVID-19 vaccine for every 1,000 residents of each race. Other races and Hispanic/Latino/a/e/x populations included in initial logistic regression models but excluded by model-fitting and selection processes.

| Allocation program | $\beta$ | SE | 95% CI | | $p$ |
| --- | --- | --- | --- | --- | --- |
|  |  |  | LL | UL |  |
| Pfizer-retail |  |  |  |  |  |
| White | .150 | .075 | 0.003 | 0.297 | .045 |
| Moderna-retail |  |  |  |  |  |
| Black | .219 | .060 | 0.101 | 0.337 | < .001 |
| White | .136 | .030 | 0.077 | 0.195 | < .001 |
| DoH |  |  |  |  |  |
| Black | -.270 | .081 | -0.429 | -0.111 | < .001 |
| White | .132 | .026 | 0.081 | 0.183 | < .001 |

**Figure 1.**
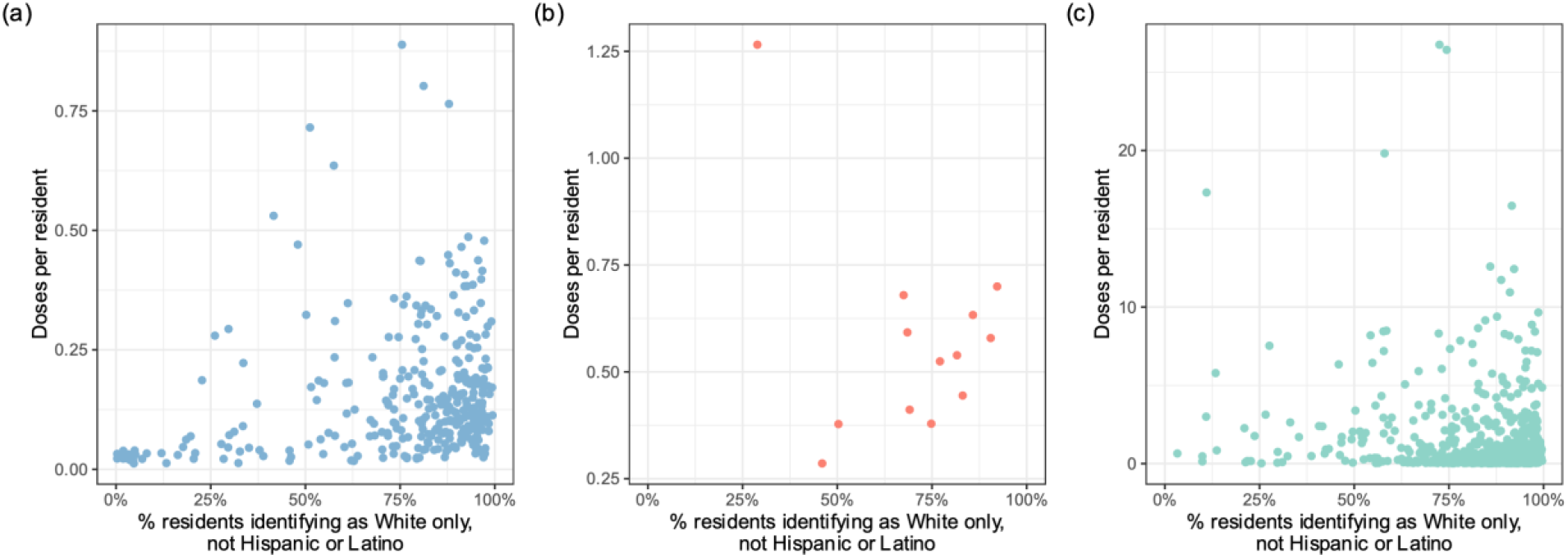
Doses-per-person by neighborhood Whiteness. (a) Moderna-retail shipments, allocated by pharmacy chains to retail locations. (b) Pfizer-retail shipments, allocated by pharmacy chains to retail locations. (c) DoH shipments, allocated by the State of Pennsylvania. Neighborhoods receiving zero doses and high-leverage data points (Cook’s *d* >4/*n*) removed for visualization.

Neighborhoods with more White residents were more likely to receive any vaccine shipments, and such neighborhoods also tended to receive more vaccine doses in those shipments. Consequently, White people had an average of 102.3% more Pfizer-retail doses shipped to their neighborhoods (*M*_*DOSES*_ = 39.7, *SD =* 0.09) than Black people did (*M*_*DOSES*_ = 19.6, *SD =* 0.06). This difference was statistically significant (*t* = 3.45, p < .001).

### Moderna-retail statistical tests

A neighborhood’s odds of receiving any vaccine via Moderna-retail partnerships increased by 15.0% for every 1,000 White residents (*OR* = 1.15) and by 25.1% (*OR* = 1.25) for every 1,000 Black residents. Neighborhoods receiving vaccine received an additional 134 additional doses for every 1,000 White residents, but only 51 doses per 1,000 Black residents (Table 3). Additional residents of other races or ethnicities had no additional effect on either of these outcomes.

**Table 3:** Expected doses per 1,000 neighborhood residents of each race. Sample sizes determined by number of neighborhoods included in each program. Other races and Hispanic/Latino/a/e/x populations included in initial linear regression models but excluded by model-fitting process.

| Allocation program | $R^2$ | $F$ | Estimate | $SE$ | $p$ |
| --- | --- | --- | --- | --- | --- |
| **Pfizer-retail** ($n = 48$) | 0.64 | 85.6 | | | < .001 |
| White |  |  | 599.4 | 64.8 | < .001 |
| **Moderna-retail** ($n = 455$) | 0.60 | 347 | | | < .001 |
| White |  |  | 133.6 | 5.3 | < .001 |
| Black |  |  | 51.2 | 19.2 | .008 |
| **DoH** ($n = 672$) | 0.14 | 52.5 | | | < .001 |
| White |  |  | 1474.9 | 224.7 | < .001 |
| Black |  |  | 8183.0 | 1649.0 | < .001 |

Black population had a slightly stronger effect on the odds of a neighborhood receiving any Moderna-retail vaccine than White population did. However, White population had a much stronger effect than Black population on the *quantity* of doses shipped to such neighborhoods. As a result, the average White person in Pennsylvania had 60.3% more Moderna-retail doses (*M*_*DOSES*_ = 93.1, *SD =* 0.07) shipped to their neighborhood than the average Black person did (*M*_*DOSES*_= 57.2, *SD* = 0.06). This difference was statistically significant (*t* = 6.38, *p* < .001).

### DoH statistical tests

The odds of DoH shipping any vaccine to a neighborhood increased by 114.1% for every 1,000 White residents (*OR* = 1.14) but decreased by 13.3% for every 1,000 Black residents (*OR* = 0.76). Neighborhoods receiving DoH shipments—which were often intended for subsequent reallocation to multiple points of service^29^—could expect 1,475 doses per 1,000 White residents and 8,183 doses per 1,000 Black residents.

DoH was more likely to send at least some vaccine to neighborhoods with larger White populations and less likely to go to neighborhoods with larger black populations. However, the number of doses sent to these neighborhoods (Figure 3c) tended to decrease as White population increased. The net result of these effects was a 16.6% advantage in average number of neighborhood doses for White (*M*_*DOSES*_ = 1913) and Black (*M*_*DOSES*_ = 1641) Pennsylvanians, though this difference was not significant according to standard criteria (*t* = 1.19, *p* = .13).

## Discussion

These findings support the view that Black-White racial disparities in COVID-19 vaccination rates during Phase 1 of Pennsylvania’s vaccination plan reflect racial inequities in COVID-19 vaccine allocation.

All three allocation programs provided greater neighborhood-level access to COVID-19 vaccines for White people than for Black people. Strikingly, these inequitable outcomes (*H*_3_) persisted across allocation programs, even though no two programs followed the same racialized pattern in both shipment of any vaccine (*H*_1_) and quantity of doses in those shipments (*H*_2_). Note that because Black and White populations were the only significant factors in our best-fitting regression models, we limit our discussion here to these two populations.

Pfizer-retail was more likely to ship *some vaccine* to Whiter neighborhoods, and for all neighborhoods that did receive shipments, it sent *more doses* to Whiter neighborhoods. Consequently, White people (46.3) had an average of 135% more Phase 1 Pfizer-retail doses in their neighborhood than Black people did (19.6). Moderna-retail was actually more likely to ship *some vaccine* to neighborhoods with larger Black populations, but it shipped *more doses* to Whiter neighborhoods. The net effect of this was a 62.8% advantage in the average number of neighborhood-level doses this program provided White people (93.1) as compared to Black people (57.2). DoH did roughly the opposite of Moderna-retail, disproportionately sending at least *some vaccine* to Whiter neighborhoods but sending far *fewer doses* to neighborhoods with larger Black populations. Though the racial disparity in quantity of neighborhood-level doses was smallest for DoH, that program still conferred a 16.5% advantage to White Pennsylvanians (1931) over Black Pennsylvanians (1641).

Collectively, these findings also suggest that public-private partnerships might produce more inequitable outcomes than state-run programs (*H*_4_). However, to determine whether this is generally the case, additional analyses of other states’ data are needed. Still, in Pennsylvania the White advantage in vaccine access was greatest when corporations with retail locations, rather than state government, determined where to send vaccine. This discrepancy is most apparent when comparing the Pfizer-retail allocations to DoH allocations. Despite federal approval to ship vaccine to any of more than 1,000 pharmacy locations in Pennsylvania (and over 40,000 locations nationwide)^32^, Pfizer-retail sent 0 of its 127,530 Pennsylvania doses to neighborhoods with fewer than 28.9% White residents. The program completely overlooked the 269 least-White neighborhoods in the state.

While the racial inequities in vaccine *allocation* we have identified are striking, we believe that they likely understate inequities in vaccine *access*. There are a number of factors that may compound the effects of vaccine allocation inequities in vaccine access inequities, and we have not attempted to study them here. In addition to having less neighborhood-level access to vaccine, Black Pennsylvanians may have been less able to obtain vaccine outside their own neighborhoods, given racial disparities in transportation access^43^, work commute time^44^, and internet access^45^. Even for those living in neighborhoods with ample vaccine supply, we might expect racial inequities in the opportunity to be vaccinated, given that White people were up to 40% more likely than Black people to be eligible for telework during the COVID-19 pandemic^46–48^. These are critical issues for policymakers to consider for future vaccine rollouts, and the failure to address them could have deadly consequences for people of all races.

Even from a so-called “race-neutral” perspective^49,50^, where the goal is to reduce overall harm to society as a whole^51^, the racial disparities we have identified cannot be justified. The Pennsylvania COVID-19 Task Force allocation of initial doses was intended to reflect “disease epidemiology and local community factors^9^.” So why send less vaccine to areas with more Black people, given that they tended to have more face-to-face interactions during the pandemic than White people did^52,53^? Pennsylvania’s Phase 1 sought to prioritize “critical populations^29^” for vaccination. So why make it harder for Black people—who were 21.9% more likely to be employed as frontline workers during the pandemic**—**to get vaccinated^54^?

Medical ethicists and epidemiologists recognize that there might always be tradeoffs between racial equity and overall utility^25,55^, but the allocation patterns we observed appear to have thwarted both virtues. By the end of Phase One, approximately one in five doses allocated to Pennsylvania by the CDC had not been administered to anyone at all^16^.

For all three vaccine allocation programs analyzed here, race had an outsized impact on where vaccine shipments went, how many doses those shipments contained, or both. That said, it would be premature to conclude that the inequitable vaccine distribution patterns we discovered in these data were the result of racial targeted discrimination. Perhaps other differences between neighborhoods that correlate with race, such as median income^56^, led corporations to allocate more vaccine to retail pharmacy locations whose patients were most likely to make purchases while in-store for their shots. For the much larger quantity of doses allocated by DoH, the geographic distribution of hospitals, pharmacies and other semi-permanent physical structures may have made it nearly inevitable that vaccine would be shipped to the Whitest neighborhoods.^57^ We think that both explanations are quite plausible—not because we have analyzed any data to support them, but because we are aware of the persistent health effects of institutional and structural racism of this kind^8,58,59^. Future research should investigate the relationship between vaccine allocation, racial demography, and “third variables” like these.

In the meantime, policies are urgently needed to increase oversight, transparency, and racial equity into the vaccine allocation process. As of this writing, new COVID-19 variants of concern have been emerging roughly once every six months^60,61^. Pharmaceutical companies have proposed to manufacture and distribute new and improved COVID-19 vaccines as soon as this month^40,41^. And a genuine commitment to vaccine equity—both within and beyond the U.S.^22^**—** is long overdue.

## Methods

### Study design

The setting, location, time period, and data collection for this research were determined by the considerations described in the introduction.

The Pennsylvania Department of Health (*DoH)* began allocating vaccine to private medical practices, hospital systems, county and municipal governments, and other entities the week of December 14, 2020. During the week of Martin Luther King Jr. Day (January 17, 2021), Moderna began allocating shipments directly to Rite Aid and Topco pharmacies through public-private partnerships with the federal government (*Moderna-retail)*^62^. Starting the week of February 21, Pfizer initiated similar partnerships with Rite Aid, Topco, CVS, and Walmart (*Pfizer-retail)*. After the week of March 8, 2021, Moderna-retail and Pfizer-retail shipments in Pennsylvania either ceased or ceased to be reported on the DoH website. Phase One vaccinations concluded on April 12, 2021^11^.

Our primary outcome measures were neighborhoods’ status with respect to receiving any vaccine shipments (a binary variable) and total doses received (a continuous variable). All available data concerning the quantities and destinations of COVID-19 vaccine shipped to Pennsylvania through by DoH, Moderna-retail, and Pfizer-retail were downloaded from the Department of Health (DoH) website^42^, to which they had been posted by the state on a weekly basis.

Our predictors were racial and ethnic population subtotals within neighborhoods (defined here as census tracts). Estimates of the racial and ethnic population totals in each of Pennsylvania’s 3,218 census tracts were obtained from the 2015-2019 American Community Survey 5-Year Estimate Data Profiles^21,63^. Since geodemographic data in the U.S. reflect historical and current practices of hypodescent (“the one-drop rule”)^64^, we drew on estimates of populations identifying as “One race, [Race], not Hispanic or Latino,” “Multiracial, not Hispanic or Latino,” and “Hispanic or Latino.” We recognize the critical limitations of this simplistic and non-exhaustive approach and discuss this elsewhere^65,66^.

### Data Processing

After resolving formatting inconsistencies across public datasets, and between those datasets and USPS address standards, we identified each vaccine shipment with the census tract of its destination. To achieve this, all shipment addresses were uploaded to the Census Bureau’s Geocoding Services Web Application Interface (API) for batched geographic identification. Addresses that were not initially geoidentified successfully were then mapped to their geographic coordinates (geocoded) via the Google Maps API. These coordinates were then uploaded to the Census Bureau’s geocoding API for batched geographic identification. All addresses that remained unidentified due to absent or errant city or ZIP code data (0.28%) were manually assigned to the PA census tracts of identical street addresses elsewhere in our dataset.

Dose quantities were missing for some Moderna-retail shipments during the first and third weeks of that program. These were imputed as 100 doses, which was both the modal and minimal allocation quantity for Moderna-retail shipments during that time. Because of heterogeneity between the three allocation programs and their data (concerning quantities shipped, time period of operation, and personnel involved), Moderna-retail, Pfizer-retail, and DoH allocations were analyzed separately for all inferential statistics. Descriptive statistics, which we include in order to provide a snapshot of the outcomes of these allocation programs from the perspective of Pennsylvania residents, are reported statewide rather than exclusively by allocation program.

### Statistical analyses

The allocation of any vaccine to a neighborhood depends on the prior availability of suitable vaccination sites and cold storage facilities. In contrast, the quantity of doses shipped to neighborhoods in which these are sites and facilities are available is somewhat more discretionary. Since the processes leading to these two outcomes are distinct (one taking place primarily pre-pandemic, the other during the pandemic), we ran three analyses for each of the three datasets.

First, we ran logistic regressions to estimate the effects of racial and ethnic population subtotals on receipt of any vaccine. We selected the best-fitting models for each allocation program using the approach advocated by Hosmer and Lemeshow^67^, and their final selection was validated in all cases using the Akaike information criterion.

Second, we conducted linear regressions to estimate the expected number of doses shipped to neighborhoods in each allocation program as a function of racial and ethnic population subtotals. Best-fitting models were selecting using the approach described in Cohen et al^68^.

Third, we estimated the net impact of these two effects (on *any doses* and on *quantity of doses, if any*) to White and Black populations’ relative opportunity to be vaccinated. For these estimates, we also report the average number of doses to which Black and White Pennsylvanians had neighborhood-level access. In line with existing research into geodemographic health inequities^69,70^, we calculated these averages as the weighted mean neighborhood doses for the racial subpopulation across all 3,218 neighborhoods in our dataset:

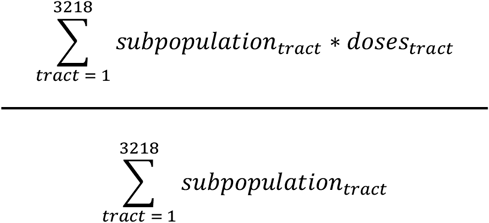

Weighted variance was calculated with this same approach to provide standard deviations and facilitate reporting of population-weighted t-tests^71^.

## Data Availability

All data produced in the present study are available upon reasonable request to the authors

## Acknowledgements

We thank Neda Khoshkhoo, Elaine Nsoesie, Yanique Redwood, and Caty Taborda for their valuable feedback on earlier drafts. We also thank Piranavakumar Kandaswamy for assisting with data validation and code management.

## Notes

### Competing Interest Statement

The authors have declared no competing interest.

### Funding Statement

This study did not receive any funding

